# Burden and risk factors for Achilles tendinopathy in the military population from 2006 to 2015. A retrospective cohort study

**DOI:** 10.1101/2023.02.27.23286521

**Authors:** Christopher K Sullivan, Cory F. Janney, John J. Fraser

**Affiliations:** Department of Orthopedic Surgery, Naval Medical Center San Diego, 34800 Bob Wilson Dr, San Diego, CA 92134, USA; Operational Readiness & Health Directorate, Naval Health Research Center, 140 Sylvester Road, San Diego, CA 92106-3521, USA

## Abstract

**Background:** Ankle-foot conditions are ubiquitous in the US Military. The objective of this study was to evaluate the burden and associated factors of Achilles tendinopathy (AT).

**Methods:** The Defense Medical Epidemiology Database was used to identify all diagnosed AT in military personnel from 2006 to 2015. Prevalence of AT was calculated and compared by year, service branch, and military rank. Unadjusted and adjusted assessment of injury risk were calculated.

**Results:** Officers incurred 37,939 episodes at a prevalence of 17.65 per 1000 (male officers: 18.20 per 1000; female officers: 14.80 per 1000). Among enlisted personnel, there were 116,122 episodes of AT that occurred in 12.22 per 1000 (male enlisted: 12.07 per 1000; female enlisted: 13.22 per 1000). All officer specialties had significantly higher risk of AT episodes compared to the ground and naval gunfire officers (PR: 1.04-1.43), with aviation demonstrating a significant protective effect (PR: 0.65). Among enlisted specialties, maritime/naval specialties had reduced risk (PR: 0.82), with all specialties (except aviation) having increased risk of AT compared to infantry (PR: 1.07-1.71). There were multiple associated factors identified, to include sex, age, rank, military occupation, and service branch.

**Conclusions:** AT was ubiquitous in the US military, with a progressive increase in prevalence during the study epoch. There were multiple associated factors identified, to include sex, age, rank, military occupation, and service branch. These findings highlight both the need for prophylactic interventions and identification of the populations with the greatest need.

## INTRODUCTION

Achilles tendinopathy (AT) is common in both military and civilian populations that affect both function and health-related quality of life. AT is characterized by persistent tendon pain and loss of function related to mechanical loading.^1^ and is classified as being insertional or non-insertional (mid-portion).^2^ The incidence of AT has been estimated to be 2.16 per 100000 person-years in the general population, although the actual burden is likely much higher as many people would not seek medical treatment for this condition.^3^ This repetitive microtraumatic injury contributes impairments in the ability to generate force and power during propulsion, jumping, and landing, which preclude activities such as walking, marching/hiking, running, and obstacle negotiation during recreation and when performing occupational duties.

The burden of AT not only affects the individual with the condition, but also the organization and society in which the individual belongs. The combined indirect and direct costs of the condition estimated to be $991 USD per patient due to resources required for providing care, lost work productivity, and absenteeism.^4^ Since AT in military members can result in limited duty, protracted and costly medical management, lost working time, and decreased military readiness, identification of the associated factors that may contribute to this injury may help to prioritize targeted prophylactic interventions.

In a recent systematic review of risk factors associated with AT in the general population, alcohol use, training during cold weather, prior lower limb tendinopathy or fracture, use of fluoroquinolone antibiotics, decreased plantar flexor strength, decreased forward progression of propulsion and a more lateral plantar pressure progression during the stance phase of gait have been identified as important.^5^ In the military population, number of deployments, moderate alcohol use, body mass, and increased age have been identified as salient factors for AT.^6^ What is still unknown is how military occupational category, which are highly heterogenous and carry their own unique hazards, affects these outcomes. While there is conflicting evidence pertaining to the role of sex in the development of AT,^5^ assessment of this factor within military occupational category is also warranted. Therefore, the purpose of this study was to determine the prevalence of AT in the military population with consideration of military occupation and identify salient risk factors that may contribute to this condition.

## METHODS

A population-based epidemiological retrospective cohort study of all service members in the US Armed Forces was performed assessing sex, military occupation, rank, branch, and year on AT prevalence from 2006-2015. The Defense Medical Epidemiological Database ([DMED], Defense Health Agency, Falls Church, VA, https://bit.ly/DHADMEDv5) was utilized to identify relevant healthcare encounters. This database provides aggregated data for International Classification of Diseases, Ninth Revision (ICD-9-CM) codes and de-identified patient characteristics, including sex, categories of military occupations, and branch of service for all active duty and reserve military service members. The database is HIPAA compliant, does not include any personal identifiable or personal health information, and has been used previously for the epidemiological study of ankle-foot injury in the military.^7,8^ This study was approved as non–human-subjects research by the Institutional Review Board at the Naval Health Research Center (NHRC.2020.0207-NHSR). The Strengthening the Reporting of Observational Studies in Epidemiology (STROBE) guidelines^9^ were used to guide reporting.

The database was queried for the number of distinct medical encounters for a primary diagnosis of AT (ICD-9-CM code 726.71) from 2006 to 2015. Prevalence calculations of patients diagnosed with AT were conducted for male and female military members, enlisted and officers, in each service branch (Army, Navy, Marine Corps, and Air Force) and occupational category.^7,8^ Prevalence ratio (PR) point estimates and 95% confidence intervals (CIs), risk difference point estimates, attributable risk (AR), number needed to harm (NNH), and chi-square statistics were calculated in the assessment of sex and occupation category using Microsoft Excel for Mac 2016 (Microsoft Corp., Redmond, WA) and a custom epidemiological spreadsheet.^10^ In the unadjusted assessment of sex and occupation on AT risk, male service members, enlisted infantry, and ground/naval gunfire officer groups were used as the reference categories.

Due to overdispersion of the data, a negative binomial model was employed over a Poisson model for the adjusted assessment of female sex (male reference), branch (US Army reference), officer rank (enlisted reference), and year (2006 reference) on the risk of ATR.^11^ Since there were excess zeros noted in the data, with many demographic categories not having any reported outcomes of interest (especially among the smaller subpopulations), assessment of convergence between unadjusted and adjusted (hurdle) negative binomial regression models were performed. In the hurdle negative binomial regression model, a logistic link is employed to remove demographic categories that contributed no episodes of ATR from the final count model. By doing so, excess zeros that contribute to skewed point estimates, standard errors, and overdispersion are parsed in the truncated regression model.^11^ Results of hurdle negative binomial are reported using calculations of the predictors regressed on count data (count model assessing the prevalence of the outcome), as well as a linked logistic regression (zero model which assessed the probability within demographic categories of not having the outcome). Both unadjusted and adjusted models are reported in the supplemental material. The regression analyses were performed using the ‘PSCL’ (version 1.5) and ‘MASS’ (version 7.3-58.1) packages on R (version 3.5.1, The R Foundation for Statistical Computing, Vienna, Austria). The level of significance was *p* < 0.05 for all analyses. PR point estimates were considered statistically significant if CIs did not cross the 1.00 threshold. Convergence of *p*-values, effect sizes, and 95% confidence intervals were considered when evaluating significant findings.

## RESULTS

**Supplemental tables 1** and **2** detail the counts and prevalence of AT in officer and enlisted personnel. In the study epoch, officers incurred 37,939 episodes at a prevalence of 17.65 per 1000 (male officers: 18.20 per 1000; female officers: 14.80 per 1000). Among enlisted personnel, there were 116,122 episodes of AT that occurred at a prevalence of 12.22 per 1000 person (male enlisted: 12.07 per 1000; female enlisted: 13.22 per 1000). **Table 1** details the univariate factor of sex within occupational specialty on AT episodes. In the officer community, female officers had significantly lower risk of AT compared to their male counterparts in all occupations (PR: 0.52-0.81). Female enlisted had a significantly higher risk in the maintenance and maritime/naval specialties and in the aggregate of all occupations (PR: 1.04-1.33). Female sex was a protective factor in the administrative and logistic enlisted specialties (PR: 0.94-0.95).

**Table 1.** Assessment of Achilles tendinopathy risk by sex within occupation in members of the US Armed Forces, 2006-2015

| **Occupational Specialty** | **PR** | **95% CI** | **Risk difference (per 1000)** | **AR %** | **p-value** | **NNH** |
| --- | --- | --- | --- | --- | --- | --- |
| <u>Enlisted Specialties</u> |  |  |  |  |  |  |
| Artillery/Gunnery | 1.04 | (0.89-1.22) | 0.5 | 3.7 | 0.64 | 2075 |
| Aviation | 0.93 | (0.72-1.21) | -0.6 | -7.1 | 0.61 | -1792 |
| Combat Engineers | 0.96 | (0.66-1.39) | -0.4 | -4.6 | 0.81 | -2310 |
| Maintenance | **1.08** | **(1.04-1.11)** | **0.8** | **7.0** | **<0.001** | **1202** |
| Administration, Intelligence, & Communication | **0.94** | **(0.91-0.96)** | **-1.0** | **-6.9** | **<0.001** | **-1010** |
| Logistics | **0.95** | **(0.91-0.99)** | **-0.6** | **-4.9** | **0.02** | **-1651** |
| Maritime/ Naval Specialties | **1.33** | **(1.15-1.55)** | **2.3** | **25** | **<0.001** | **434** |
| Enlisted Total | **1.04** | **(1.02-1.06)** | **1.2** | **8.7** | **<0.001** | **866** |
| <u>Officer Specialties</u> |  |  |  |  |  |  |
| Ground / Naval Gunfire | **0.52** | **(0.43-0.63)** | **-8.1** | **-91.9** | **<0.001** | **-123** |
| Aviation | **0.63** | **(0.53-0.74)** | **-4.1** | **-58.9** | **<0.001** | **-243** |
| Engineering & Maintenance | **0.84** | **(0.77-0.91)** | **-3.1** | **-18.9** | **<0.001** | **-318** |
| Administration | **0.76** | **(0.70-0.82)** | **-6.0** | **-31.4** | **<0.001** | **-168** |
| Operations & Intelligence | **0.78** | **(0.71-0.85)** | **-4.6** | **-29.0** | **<0.001** | **-218** |
| Logistics | **0.78** | **(0.72-0.84)** | **-5.6** | **-28.7** | **<0.001** | **-180** |
| Services | **0.70** | **(0.67-0.74)** | **-5.7** | **-42.2** | **<0.001** | **-176** |
| Officer Total | **0.81** | **(0.79-0.84)** | **-3.4** | **-22.9** | **<0.001** | **-295** |

Female service members referenced to male members. PR, prevalence ratio; CI, confidence interval; AR, attributable risk; NNH, number needed to harm. Bolded values indicate statistical significance.

In the assessment of occupational exposure, all but one officer specialties had significantly higher risk of AT episodes compared to the ground and naval gunfire officers (PR: 1.04-1.43) (**Table 2**). Aviation was the only officer community that demonstrated a significant protective effect (PR: 0.65). Among enlisted specialties, maritime/naval specialties had reduced risk (PR: 0.82), with all specialties (except aviation) having increased risk of AT compared to infantry (PR: 1.07-1.71). There were no significant difference in risk between aviation specialties and infantry.

**Table 2.**
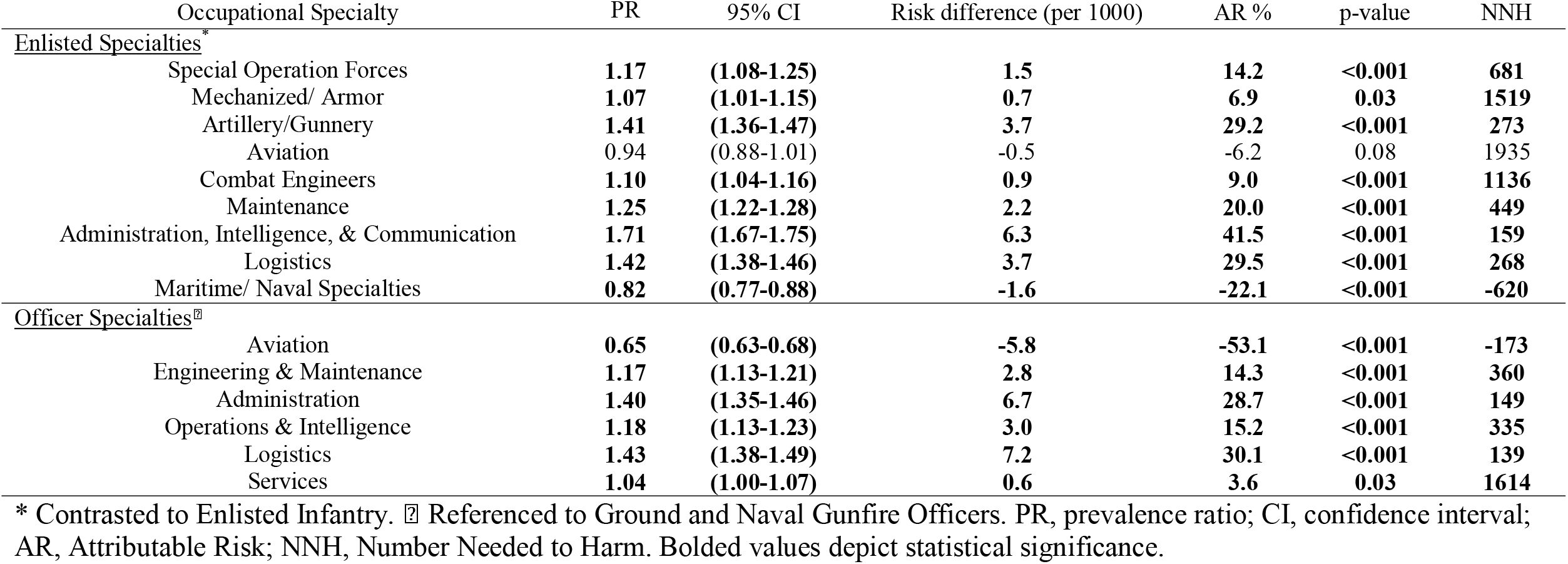
Assessment of Achilles tendinopathy risk by occupation in the US Armed Forces, 2006-2015

| Occupational Specialty | PR | 95% CI | Risk difference (per 1000) | AR % | p-value | NNH |
| --- | --- | --- | --- | --- | --- | --- |
| <u>Enlisted Specialties*</u> |  |  |  |  |  |  |
| Special Operation Forces | **1.17** | **(1.08-1.25)** | **1.5** | **14.2** | **<0.001** | **681** |
| Mechanized/ Armor | **1.07** | **(1.01-1.15)** | **0.7** | **6.9** | **0.03** | **1519** |
| Artillery/Gunnery | **1.41** | **(1.36-1.47)** | **3.7** | **29.2** | **<0.001** | **273** |
| Aviation | 0.94 | (0.88-1.01) | -0.5 | -6.2 | 0.08 | 1935 |
| Combat Engineers | **1.10** | **(1.04-1.16)** | **0.9** | **9.0** | **<0.001** | **1136** |
| Maintenance | **1.25** | **(1.22-1.28)** | **2.2** | **20.0** | **<0.001** | **449** |
| Administration, Intelligence, & Communication | **1.71** | **(1.67-1.75)** | **6.3** | **41.5** | **<0.001** | **159** |
| Logistics | **1.42** | **(1.38-1.46)** | **3.7** | **29.5** | **<0.001** | **268** |
| Maritime/ Naval Specialties | **0.82** | **(0.77-0.88)** | **-1.6** | **-22.1** | **<0.001** | **-620** |
| <u>Officer Specialties<sup>‡</sup></u> |  |  |  |  |  |  |
| Aviation | **0.65** | **(0.63-0.68)** | **-5.8** | **-53.1** | **<0.001** | **-173** |
| Engineering & Maintenance | **1.17** | **(1.13-1.21)** | **2.8** | **14.3** | **<0.001** | **360** |
| Administration | **1.40** | **(1.35-1.46)** | **6.7** | **28.7** | **<0.001** | **149** |
| Operations & Intelligence | **1.18** | **(1.13-1.23)** | **3.0** | **15.2** | **<0.001** | **335** |
| Logistics | **1.43** | **(1.38-1.49)** | **7.2** | **30.1** | **<0.001** | **139** |
| Services | **1.04** | **(1.00-1.07)** | **0.6** | **3.6** | **0.03** | **1614** |

\* Contrasted to Enlisted Infantry. <sup>‡</sup> Referenced to Ground and Naval Gunfire Officers. PR, prevalence ratio; CI, confidence interval; AR, Attributable Risk; NNH, Number Needed to Harm. Bolded values depict statistical significance.

Table 3 details the results of the hurdle (adjusting for zero-deflation) and standard multivariable negative binomial regression models. In both assessments, female sex and ages >30 were salient factors once rank, branch, and year were controlled. Junior officers had significantly lower risk compared to junior enlisted and service in the Army had the greatest risk for AT episodes. The frequency of AT episodes were significantly higher from 2010 to 2016 compared to 2006.

**Table 3.** Results of the adjusted and unadjusted negative binomial regressions assessing sex, age, rank, service branch, and year on prevalence of Achilles tendinopathy in the US Armed Forces, 2006-2015.

|  |  | Hurdle Negative Binomial Model<br>(adjusted for zero-inflation) |  |  | Unadjusted Negative Binomial |  |  |
| --- | --- | --- | --- | --- | --- | --- | --- |
|  |  | 95% CI |  |  | 95% CI |  |  |
|  |  | PR | LL | UL | PR | LL | UL |
| **Sex:** Females |  | **1.21** | **1.13** | **1.29** | **1.16** | **1.06** | **1.26** |
| **Age Range:** 20-29 |  | **0.80** | **0.69** | **0.93** | **0.73** | **0.60** | **0.88** |
| 30-39 |  | **1.54** | **1.34** | **1.78** | **1.29** | **1.08** | **1.55** |
| 40+ |  | **3.06** | **2.64** | **3.54** | **2.34** | **1.93** | **2.82** |
| **Rank:** |  |  |  |  |  |  |  |
| Senior Enlisted |  | **0.78** | **0.71** | **0.87** | 0.93 | 0.82 | 1.05 |
| Junior Officers |  | **0.68** | **0.61** | **0.75** | **0.74** | **0.65** | **0.83** |
| Senior Officers |  | **0.80** | **0.72** | **0.90** | 0.94 | 0.82 | 1.09 |
| **Branch:** US Navy |  | **0.55** | **0.50** | **0.60** | **0.51** | **0.45** | **0.58** |
| US Marine Corps |  | **0.89** | **0.81** | **0.97** | **0.74** | **0.65** | **0.83** |
| US Air Force |  | **0.91** | **0.83** | **1.00** | **0.87** | **0.77** | **0.98** |
| **Year:** 2007 |  | 1.03 | 0.88 | 1.20 | 1.07 | 0.88 | 1.30 |
| 2008 |  | 0.97 | 0.83 | 1.14 | 0.95 | 0.78 | 1.15 |
| 2009 |  | 1.04 | 0.89 | 1.22 | 1.06 | 0.87 | 1.29 |
| 2010 |  | **1.28** | **1.10** | **1.49** | **1.36** | **1.12** | **1.65** |
| 2011 |  | **1.31** | **1.12** | **1.52** | **1.37** | **1.13** | **1.67** |
| 2012 |  | **1.43** | **1.23** | **1.67** | **1.49** | **1.23** | **1.81** |
| 2013 |  | **1.58** | **1.36** | **1.84** | **1.67** | **1.37** | **2.02** |
| 2014 |  | **1.56** | **1.34** | **1.81** | **1.60** | **1.32** | **1.94** |
| 2015 |  | **1.63** | **1.40** | **1.90** | **1.69** | **1.40** | **2.05** |
|  |  | Log-Likelihood Ratio:<br>-3025 on 41 *Df* |  |  | Log-Likelihood Ratio:<br>-6583 on 41 *Df* |  |  |

Bolded figures depict statistical significance ( $p < .05$ ); PR, prevalence ratio; CI, confidence interval. The zero model employed a binomial regression with logit link. In the second step, the count model employed a truncated negative binomial regression (omitting zero counts) with log link. Contrasts: Sex [Male]; Age [18-19]; Rank [Junior Enlisted]; Branch [US Army]; Year [2006]

## DISCUSSION

The primary findings of this study were that AT was ubiquitous in the US military with multiple salient factors identified, to include sex, age, rank, military occupation, and service branch. In the assessment of outcomes over time, the number of care episodes were incrementally higher in the latter years of the study. Based on the growing burden of AT in the US military observed in this study, these findings highlight both the need for prophylactic interventions and identification of the populations who can most greatly benefit from such interventions.

There are few plausible reasons why AT was more common in the latter half of this study. The operational tempo of military operations earlier in the study epoch likely influenced the degree of exposure to potentially injurious loads that may contribute to AT. The US military was involved in contingency operations in both Iraq and Afghanistan, with drawdowns occurring in both theaters beginning in 2011. It is plausible that cumulative exposure to increased load carriage (due to body armor, equipment, etc.) earlier in the study may have resulted in the manifestation of repetitive microtrauma in the later years. It is also conceivable that higher operational tempo earlier in the study epoch precluded the ability to seek care, with military drawdown providing an opportunity to seek care for more chronic conditions that were put off due to prioritization of the mission.^12^ Another possible explanation of increased prevalence later in the study epoch may be associated with force shaping requirements, with the necessity to open the recruitment aperture in order to meet mission requirements.^12^ There is evidence for a progressive increase in body mass among recruits^13^ and active military members^14^ over time, a factor that has been shown to be associated with increased AT risk.^5,6^

Occupation was identified as a salient factor for AT, a finding that has been previously reported in military populations.^6^ Disparities in burden between occupations may be a function of the types of exposures and hazards unique to that vocation encountered during operations. In addition, there are likely social factors that either foster or preclude the ability to seek care, especially when absence from the unit would affect the ability to meet mission requirements.^12^ Cultural differences between occupations may also explain a servicemember chooses to seek care, or the degree to which a supervisor is willing to support and encourage medical care-seeking. This supposition may also explain why the greatest risk was observed in office-based occupations compared to combat arms. Military members working in office-based occupations are frequently in greater proximity to medical services, which provides greater access and reduces the time absent from work when care is needed. Lastly, it is plausible that perceptual differences between occupations about injury and the return on investment of medical care-seeking may also drive care-seeking behaviors.^12^ These factors may also explain why there significant differences observed between military branches.

Older age was an especially salient factor for AT, with the strongest and largest magnitude effects observed for this variable. While evidence for age as a factor is mixed across populations,^5^ it appears younger age is a protective factor in military populations.^6^ From a physiological perspective, changes in elasticity and vascularity tends to decrease with age, with a curtailed ability to heal following repetitive microtrauma.^15^ Stiffness also may play a role, with women generally demonstrating higher tendon compliance.^16^ This may explain why women tend to have lower prevalence of rupture, but higher prevalence of AT.^16^ It is also plausible that these findings is tied to the retirement benefit. Military members in their 40s and 50s are retirement eligible, which would foster increased care-seeking for management of chronic AT symptoms that were put off earlier in their career. It is plausible that increased body mass in older military members was a salient covariate, since this subpopulation has been shown in prior research to contribute the greatest burden of obsesity.^14^ This supposition would require substantiation in future research.

This study demonstrates that AT is an increasing threat to medical readiness of the armed forces and a substantial burden to the Military Health System. In a resource constrained environment, these results can be used to provide targeted prevention programs to the communities with the greatest risk. Furthermore, this study can assist military leaders and medical logisticians by providing greater precision when planning the types of clinicians and supplies needed to address AT throughout the military. While treatment methods of AT is beyond the scope on this manuscript, clinicians are encouraged to consult the papers by Martin and colleague^17^ and Baltes and colleagues^18^ for additional information on the conservative and surgical management of this condition.

A substantial strength of this study was the DMED database allowed for the assessment of population-level differences in AT across several strata, including sex, occupation, military rank, branch of service, and year. There are also limitations of this study. Using diagnostic codes has inherent weaknesses, especially in military populations where many determinants can influence care seeking.^12^ The data used in this study only represent individuals who sought treatment for their injuries and omits individuals who self-managed their condition. Lastly, although personnel were categorized by their military occupation, a detailed task analysis to determine the specific duties and exposures that may be associated with these injuries was not possible.

## CONCLUSION

AT was ubiquitous in the US military, with a progressive increase in prevalence during the study epoch. There were multiple associated factors identified, to include sex, age, rank, military occupation, and service branch. Based on the growing burden of AT in the US military observed in this study, these findings highlight both the need for prophylactic interventions and identification of the populations who can most greatly benefit from such interventions.

## Data Availability

All data produced in the present work are contained in the manuscript.

---

**Supplemental Table 1.**
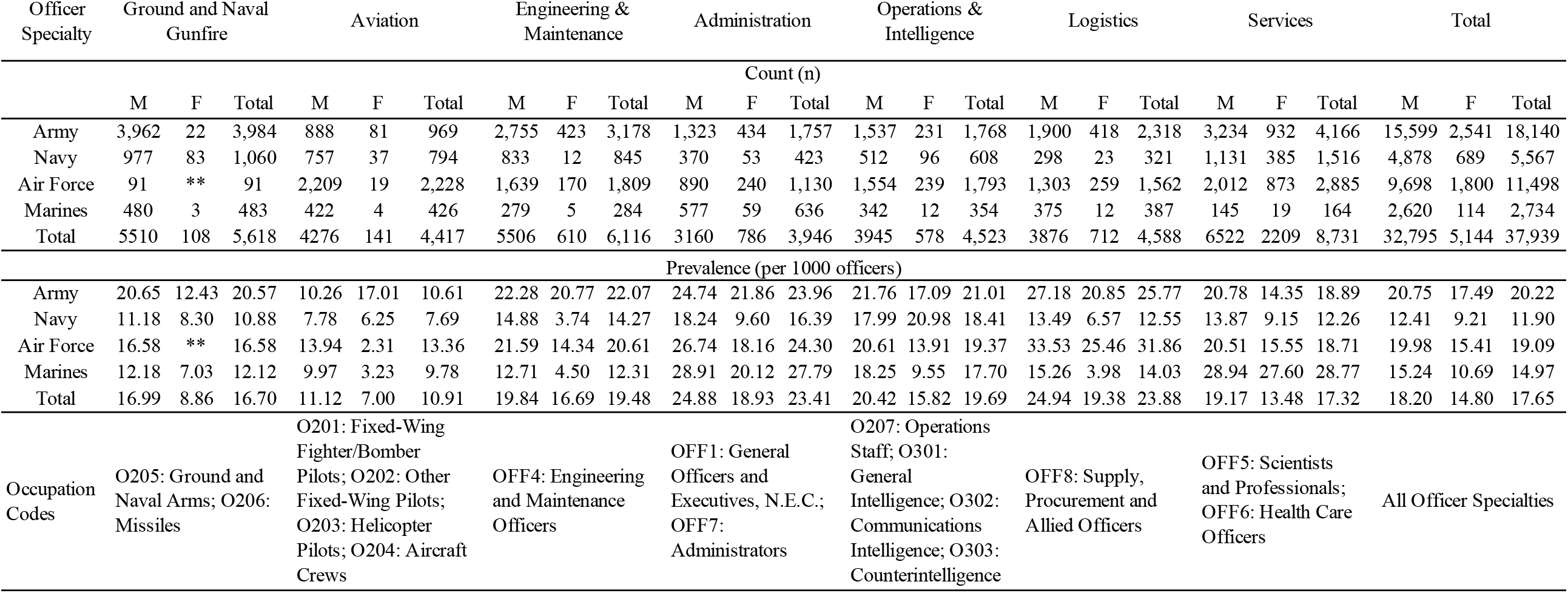
Count and prevalence of Achilles tendinopathy care episodes among officers in the US Armed Forces, 2006–2015

**Supplemental Table 2.** Count and prevalence of Achilles tendinopathy care episodes among enlisted members in the US Armed Forces, 2006–2015

| Enlisted Specialty | Special Operations Forces |  |  | Infantry |  |  | Mechanized/Armor |  |  | Artillery/Gunnery |  |  | Aviation |  |  | Combat Engineers |  |  | Maintainance |  |  | Administration, Intelligence, & Communication |  |  | Logistics |  |  | Maritime/Naval Specialties |  |  | Total |  |  |
| --- | --- | --- | --- | --- | --- | --- | --- | --- | --- | --- | --- | --- | --- | --- | --- | --- | --- | --- | --- | --- | --- | --- | --- | --- | --- | --- | --- | --- | --- | --- | --- | --- | --- |
|  | Count (n) |  |  |  |  |  |  |  |  |  |  |  |  |  |  |  |  |  |  |  |  |  |  |  |  |  |  |  |  |  |  |  |  |
|  | M | F | Total | M | F | Total | M | F | Total | M | F | Total | M | F | Total | M | F | Total | M | F | Total | M | F | Total | M | F | Total | M | F | Total | M | F | Total |
| Army | 715 | ** | 715 | 7,367 | ** | 7,367 | 923 | ** | 923 | 3,077 | 143 | 3,220 | ** | ** | ** | 1,299 | 27 | 1,326 | 13,861 | 1,586 | 15,447 | 15,436 | 4,372 | 19,808 | 7,787 | 1,576 | 9,363 | 76 | 21 | 97 | 50,541 | 7,725 | 58,266 |
| Navy | 61 | ** | 61 | ** | ** | ** | ** | ** | ** | 125 | 14 | 139 | 183 | 42 | 225 | ** | ** | ** | 6,106 | 1,122 | 7,228 | 3,515 | 890 | 4,405 | 1,315 | 242 | 1,557 | 634 | 193 | 827 | 11,939 | 2,503 | 14,442 |
| Air Force | ** | ** | ** | ** | ** | ** | ** | ** | ** | ** | ** | ** | 650 | 15 | 665 | ** | ** | ** | 13,656 | 933 | 14,589 | 10,437 | 3,486 | 13,923 | 3,525 | 644 | 4,169 | ** | ** | ** | 28,268 | 5,078 | 33,346 |
| Marines | ** | ** | ** | 1,115 | ** | 1,115 | 99 | ** | 99 | 129 | ** | 129 | 52 | 2 | 54 | 167 | 1 | 168 | 3,266 | 176 | 3,442 | 3,206 | 550 | 3,756 | 1,155 | 150 | 1,305 | ** | ** | ** | 9,189 | 879 | 10,068 |
| Total | 776 | ** | 776 | 8482 | ** | 8,482 | 1022 | ** | 1,022 | 3331 | 157 | 3,488 | 885 | 59 | 944 | 1466 | 28 | 1,494 | 36889 | 3817 | 40,706 | 32594 | 9298 | 41,892 | 13782 | 2612 | 16,394 | 710 | 214 | 924 | 99,937 | 16,185 | 116,122 |
| Prevalence (per 1000 enlisted members) |  |  |  |  |  |  |  |  |  |  |  |  |  |  |  |  |  |  |  |  |  |  |  |  |  |  |  |  |  |  |  |  |  |
| Army | 11.60 | ** | 11.60 | 11.35 | ** | 11.35 | 11.24 | ** | 11.24 | 14.93 | 17.45 | 15.03 | ** | ** | ** | 11.54 | 18.79 | 11.63 | 16.04 | 19.55 | 16.35 | 17.38 | 17.48 | 17.41 | 16.11 | 15.20 | 15.95 | 17.59 | 40.57 | 20.05 | 15.08 | 17.36 | 15.35 |
| Navy | 4.61 | ** | 4.61 | ** | ** | ** | ** | ** | ** | 5.04 | 3.62 | 4.85 | 4.17 | 9.29 | 4.65 | ** | ** | ** | 5.32 | 7.29 | 5.56 | 8.86 | 7.41 | 8.52 | 8.51 | 6.51 | 8.12 | 6.44 | 8.50 | 6.83 | 6.35 | 7.31 | 6.50 |
| Air Force | ** | ** | ** | ** | ** | ** | ** | ** | ** | ** | ** | ** | 14.97 | 6.12 | 14.50 | ** | ** | ** | 15.03 | 14.50 | 14.99 | 21.37 | 15.84 | 19.66 | 14.03 | 12.12 | 13.70 | ** | ** | ** | 16.71 | 14.94 | 16.41 |
| Marines | ** | ** | ** | 3.66 | ** | 3.66 | 3.98 | ** | 3.98 | 3.71 | ** | 3.71 | 2.91 | 3.74 | 2.94 | 4.48 | 0.64 | 4.33 | 7.81 | 8.06 | 7.82 | 9.42 | 10.21 | 9.53 | 6.40 | 8.02 | 6.55 | ** | ** | ** | 6.76 | 9.11 | 6.92 |
| Total | 10.36 | ** | 10.36 | 8.89 | ** | 8.89 | 9.55 | ** | 9.55 | 12.54 | 13.02 | 12.56 | 8.41 | 7.86 | 8.38 | 9.78 | 9.35 | 9.77 | 11.05 | 11.88 | 11.12 | 15.42 | 14.43 | 15.19 | 12.89 | 12.28 | 12.78 | 6.91 | 9.22 | 7.34 | 12.07 | 13.22 | 12.22 |
| Occupation Codes | E011: Special Forces |  |  | E010: Infantry, General |  |  | Occupation: E020: Armor and Amphibious, General |  |  | E041: Artillery and Gunnery; E042: Rocket Artillery; E043: Missile Artillery, Operating Crew |  |  | E050: Air Crew, General; E051: Pilots and Navigators |  |  | E030: Combat Engineering, General |  |  | ENL7: Craftworkers; ENL1: Electronic Equipment Repairers; ENL6: Electrical/Mechanical Equipment Repairers |  |  | ENL2: Communications and Intelligence Specialist; ENL5: Functional Support and Administration |  |  | ENL8: Service and Supply Handlers |  |  | E062: Small Boat Operators; E063: Seamanship, General; E060: Boatswains |  |  | All Enlisted Specialties |  |  |

**Supplemental Table 3.**
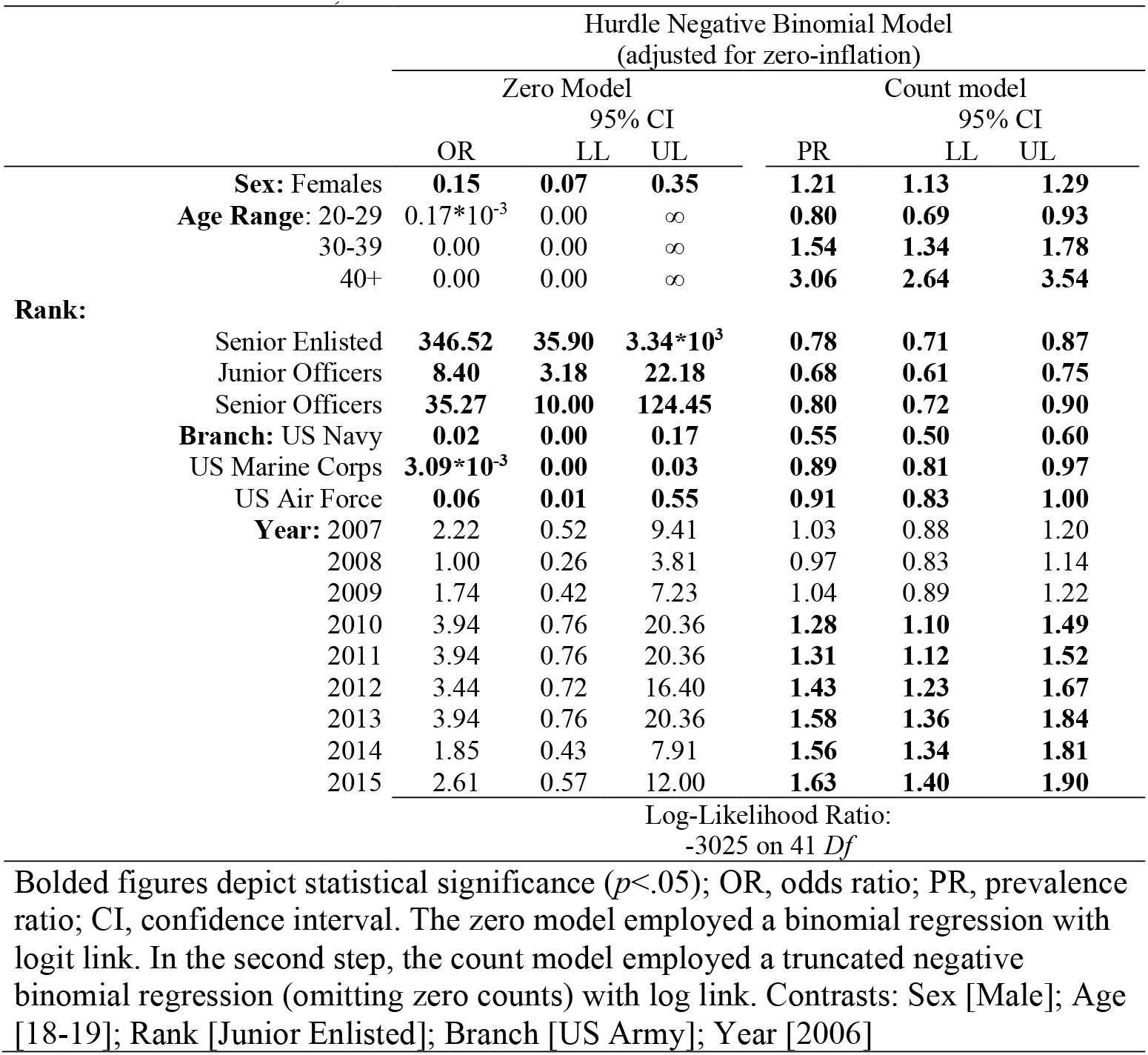
Results of the hurdle negative binomial regressions assessing sex, age, rank, service branch, and year on the prevalence of Achilles tendinopathy in the US Armed Forces, 2006-2015.

| Hurdle Negative Binomial Model<br>(adjusted for zero-inflation) |  |  |  |  |  |  |
| --- | --- | --- | --- | --- | --- | --- |
|  | Zero Model |  |  | Count model |  |  |
|  | OR | 95% CI |  | PR | 95% CI |  |
|  |  | LL | UL |  | LL | UL |
| **Sex:** Females | **0.15** | **0.07** | **0.35** | **1.21** | **1.13** | **1.29** |
| **Age Range:** 20-29 | 0.17*10<sup>-3</sup> | 0.00 | ∞ | **0.80** | **0.69** | **0.93** |
| 30-39 | 0.00 | 0.00 | ∞ | **1.54** | **1.34** | **1.78** |
| 40+ | 0.00 | 0.00 | ∞ | **3.06** | **2.64** | **3.54** |
| **Rank:** |  |  |  |  |  |  |
| Senior Enlisted | **346.52** | **35.90** | **3.34\*10<sup>3</sup>** | **0.78** | **0.71** | **0.87** |
| Junior Officers | **8.40** | **3.18** | **22.18** | **0.68** | **0.61** | **0.75** |
| Senior Officers | **35.27** | **10.00** | **124.45** | **0.80** | **0.72** | **0.90** |
| **Branch:** US Navy | **0.02** | **0.00** | **0.17** | **0.55** | **0.50** | **0.60** |
| US Marine Corps | **3.09\*10<sup>-3</sup>** | **0.00** | **0.03** | **0.89** | **0.81** | **0.97** |
| US Air Force | **0.06** | **0.01** | **0.55** | **0.91** | **0.83** | **1.00** |
| **Year:** 2007 | 2.22 | 0.52 | 9.41 | 1.03 | 0.88 | 1.20 |
| 2008 | 1.00 | 0.26 | 3.81 | 0.97 | 0.83 | 1.14 |
| 2009 | 1.74 | 0.42 | 7.23 | 1.04 | 0.89 | 1.22 |
| 2010 | 3.94 | 0.76 | 20.36 | **1.28** | **1.10** | **1.49** |
| 2011 | 3.94 | 0.76 | 20.36 | **1.31** | **1.12** | **1.52** |
| 2012 | 3.44 | 0.72 | 16.40 | **1.43** | **1.23** | **1.67** |
| 2013 | 3.94 | 0.76 | 20.36 | **1.58** | **1.36** | **1.84** |
| 2014 | 1.85 | 0.43 | 7.91 | **1.56** | **1.34** | **1.81** |
| 2015 | 2.61 | 0.57 | 12.00 | **1.63** | **1.40** | **1.90** |
| Log-Likelihood Ratio:<br>-3025 on 41 *Df* |  |  |  |  |  |  |

Bolded figures depict statistical significance ( $p < .05$ ); OR, odds ratio; PR, prevalence ratio; CI, confidence interval. The zero model employed a binomial regression with logit link. In the second step, the count model employed a truncated negative binomial regression (omitting zero counts) with log link. Contrasts: Sex [Male]; Age [18-19]; Rank [Junior Enlisted]; Branch [US Army]; Year [2006]

## Notes

### Competing Interest Statement

Dr Fraser reports grants from Congressionally Directed Medical Research Programs and the Office of Naval Research, outside of the submitted work. In addition, Dr Fraser has a patent pending for an Adaptive and Variable Stiffness Ankle Brace, U.S. Provisional Patent Application No. 63254,474.

### Funding Statement

The authors are military service members or employees of the U.S. Government. This work was prepared as part of our official duties. Title 17, U.S.C. SS105 provides that copyright protection under this title is not available for any work of the U.S. Government. Title 17, U.S.C. SS101 defines a U.S. Government work as work prepared by a military service member or employee of the U.S. Government as part of that person's official duties. The views expressed in this article are those of the authors and do not necessarily reflect the official policy or position of the Department of the Navy, Department of Defense, nor the U.S. Government.

### Author Declarations

The study protocol was approved by the Naval Health Research Center Institutional Review Board in compliance with all applicable Federal regulations governing the protection of human subjects. Research data were derived from an approved Naval Health Research Center Institutional Review Board protocol, number NHRC.2020.0207-NHSR.

